# User Testing of a Diagnostic Decision Support System with Machine-assisted Chart Review to Facilitate Clinical Genomic Diagnosis

**DOI:** 10.1101/2020.08.21.20179580

**Authors:** Alanna Kulchak Rahm, Nephi A. Walton, Lynn K. Feldman, Conner Jenkins, Troy Jenkins, Thomas N. Person, Joseph Peterson, Jonathon C. Reynolds, Peter N. Robinson, Makenzie A. Woltz, Marc S. Williams, Michael M. Segal

**Affiliations:** Genomic Medicine Institute, Geisinger, Danville, Pennsylvania, USA; Intermountain Precision Genomics, Intermountain Healthcare, St George, Utah, USA; SimulConsult Inc., Chestnut Hill MA, USA; University of Utah, Salt Lake City, UT, USA; The Jackson Laboratory for Genomic Medicine and Institute for Systems Genomics, University of Connecticut, Farmington, CT, USA

**Keywords:** health information systems, heuristic evaluation, cognitive walkthrough, Diagnostic Decision Support System (DDSS), Clinical Decision Support (CDS)

## Abstract

**Background:** There is a need in clinical genomics for systems that assist in clinical diagnosis, analysis of genomic information and periodic re-analysis of results, and can utilize information from the electronic health record to do so. Such systems should be built using the concepts of human-centered design, fit within clinical workflows, and provide solutions to priority problems.

**Methods:** We adapted a commercially available diagnostic decision support system (DDSS) to use extracted findings from a patient record and combine them with genomic variant information in the DDSS interface. Three representative patient cases were created in a simulated clinical environment for user testing. A semi-structured interview guide was created to illuminate factors relevant to human factors in CDS design and organizational implementation.

**Results:** Six individuals completed the user testing process. Tester responses were positive and noted good fit with real-world clinical genetics workflow. Technical issues related to interface, interaction, and design were minor and fixable. Testers suggested solving issues related to terminology and usability through training and infobuttons. Time savings was estimated at 30-50% and additional uses such as in-house clinical variant analysis were suggested for increase fit with workflow and to further address priority problems.

**Conclusion:** This study provides preliminary evidence for usability, workflow fit, acceptability, and implementation potential of a modified DDSS that includes machine-assisted chart review. Continued development and testing using principles from human-centered design and implementation science are necessary to improve technical functionality and acceptability for multiple stakeholders and organizational implementation potential to improve the genomic diagnosis process.

**SUMMARY:** *What is already known?:*

- There is a need in clinical genomics for tools that assist in analysis of genomic information and can do so using information from the electronic health record.
- Such tools should be easy to use, fit within clinical workflows, and provide solutions to priority problems as defined by clinician end-users.
- Natural language processing (NLP) is a useful tool to read patient records and extract findings.

*What does this paper add?:*

- We demonstrated the use of Human-centered design and implementation science principles in a simulated environment for assessment of a new version of a decision support tool prior to large-scale implementation.
- This study provides preliminary evidence that a clinical decision support tool with machine-assisted chart review is acceptable to clinical end-users, fits within the clinical workflow, and addresses perceived needs within the differential diagnosis process across all Mendelian genetic disorders.
- Terminology codes for DDSSs should have levels of granularity tuned to the sensitivity and specificity appropriate to its various functions, e.g., NLP versus chart documentation.

## INTRODUCTION

Clinical Decision Support (CDS) integrated into Electronic Health Records (EHRs) has long been considered a promising way to improve patient outcomes and decrease inefficiencies.^1-4^ It is also recognized that CDS must be designed with the user in mind, fitting the concepts of human-centered design with computer interfaces at the individual clinician level.^1 5^ Design alone, however, is insufficient to facilitate implementation. For CDS to impact care and patient outcomes, it must fit within clinician workflow and provide a solution to a priority problem for the clinician and the healthcare system.^4 6-8^

Diagnostic Decision Support Systems (DDSSs) are a key type of CDS needed in genomics to supplement a shortage of trained clinicians and address the inherent complexity of genomic diagnosis.^9 10^ This complexity arises from the heterogeneous nature of genetic diseases, the variable expression in patients, and the degree of overlap in findings (i.e., signs, symptoms, and test results) among genetic conditions, sometimes differentiated only by onset age of individual findings.^11^ Position statements and a systematic review note two new functions needed for DDSSs in genomics: (1) a cost-effective, regular approach to re-evaluation of patient cases in light of new findings or genetic knowledge, when testing does not immediately yield a diagnosis; and (2) developing machine-assisted chart review.^12 13^ Most genomic patient records are extensive with input from by multiple clinicians, such that manual review is prohibitively time-consuming; resulting in added costs from repeated or unnecessary tests and increased risk of missed information that could have facilitated timely diagnosis. Because most of the relevant information is in unstructured clinical notes, approaches such as natural language processing (NLP) are needed to automate and assist this manual process.

To address both re-evaluation and automation, we adapted a commercially available DDSS already capable of incorporating genomic sequencing data to perform automated chart review and present the information to a clinician in the form of findings obtained through structured data mining and NLP of an EHR. We then created clinical case vignettes to simulate the real-world clinical diagnostic workflow for user testing. The goal was to provide preliminary evidence of usability, perceived fit with clinical need and workflow, and potential for implementation into the real-world clinical environment.

## METHODS

### Setting

Development of the clinical case vignettes, simulated EHR environment, and user testing were conducted at Geisinger, a healthcare system in rural Pennsylvania.

### Adapting a DDSS for machine-assisted chart review of clinical findings

We adapted SimulConsult’s Genome-Phenome Analyzer, as it is the one DDSS that allows for detailed analysis of clinical information, including pertinent negatives, findings onset information, and frequency and treatability of diseases. It has also been shown to be accurate and helpful in clinical diagnosis, including interpreting genomic results.^14-16^ Described in detail elsewhere,^11 14 15^ SimulConsult correlates annotated variant call files (VCFs) with patient-specific clinical and family history information; and the underlying algorithms include age-dependent Bayesian pattern-matching and computational metrics of usefulness and pertinence. SimulConsult also generates a Patient Summary for saving interim patient findings and a customizable genomic Return of Results (RoR) report shown in previous research to be effective for facilitating standardized communication for patients and referring clinicians.^17-20^ When clinicians enter findings, the DDSS returns a ranked list of candidate diseases and suggestions of other findings to check, ranked by usefulness in narrowing the differential diagnosis in a way that accounts for cost and treatability; thus facilitating the iterative approach of information gathering in diagnosis.^21 22^ For each finding, presence (with onset age) or absence can be specified (Figure A).

**Figure A.**
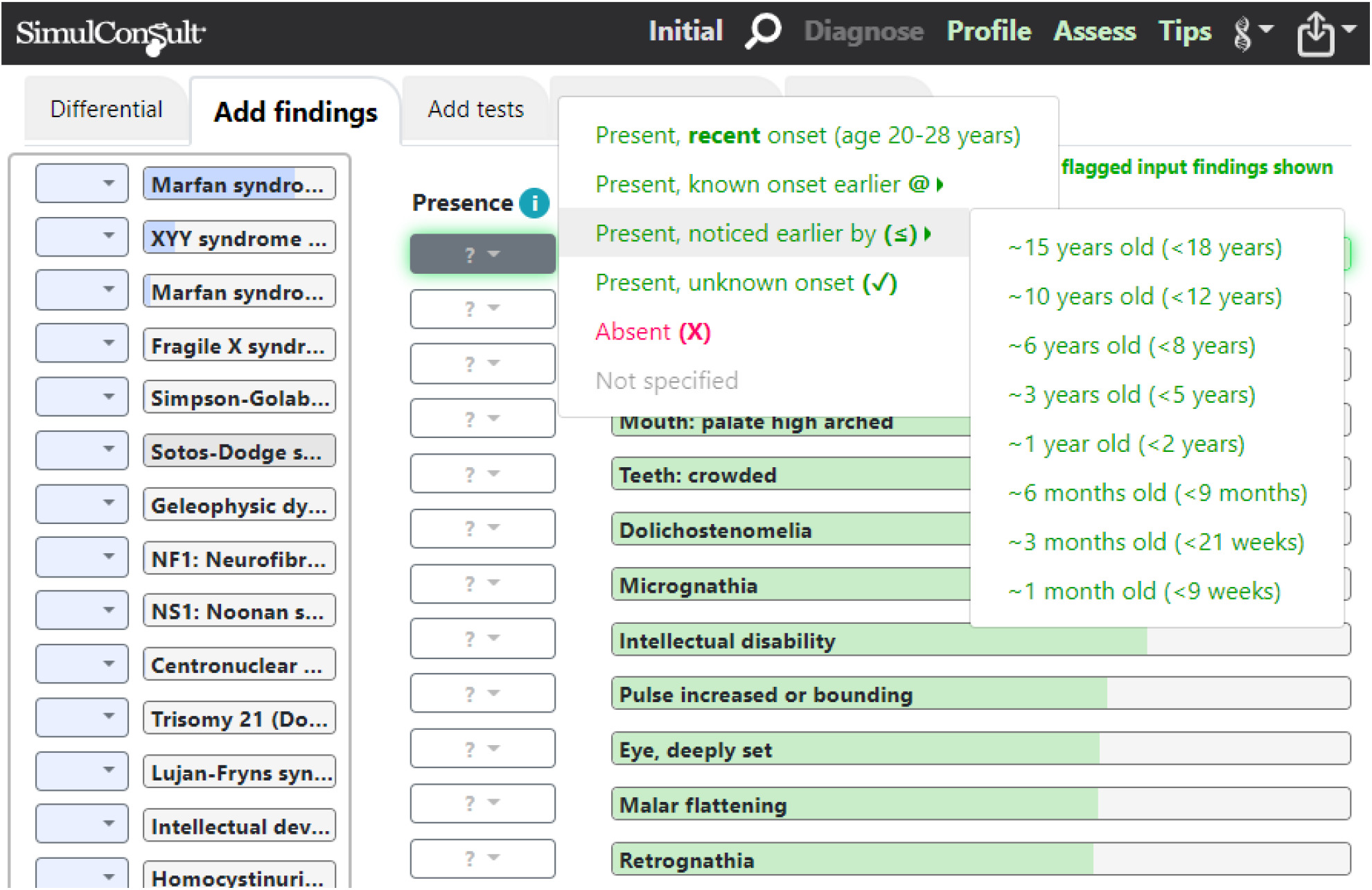
SimulConsult main interface showing ranked list of candidate diseases and guidance for entering finding presence (or absence) with onset age.

We used the Logica platform to create a simulated EHR and the cTAKES tool with the Unified Medical Language System (UMLS) module^23^ for NLP of patient notes. Steps in adaption included (1) mapping DDSS findings to Human Phenotype Ontology (HPO) and UMLS codes, including creation of hundreds of new HPO terms resulting in creation of new UMLS concepts, (2) using results from NLP analysis of EHR notes to flag “Mentions” of the findings used by the DDSS, and (3) augmenting the DDSS’s interface to present the flagged findings with contextual information needed to clinically assess the information (Table 1).

**Table 1.**
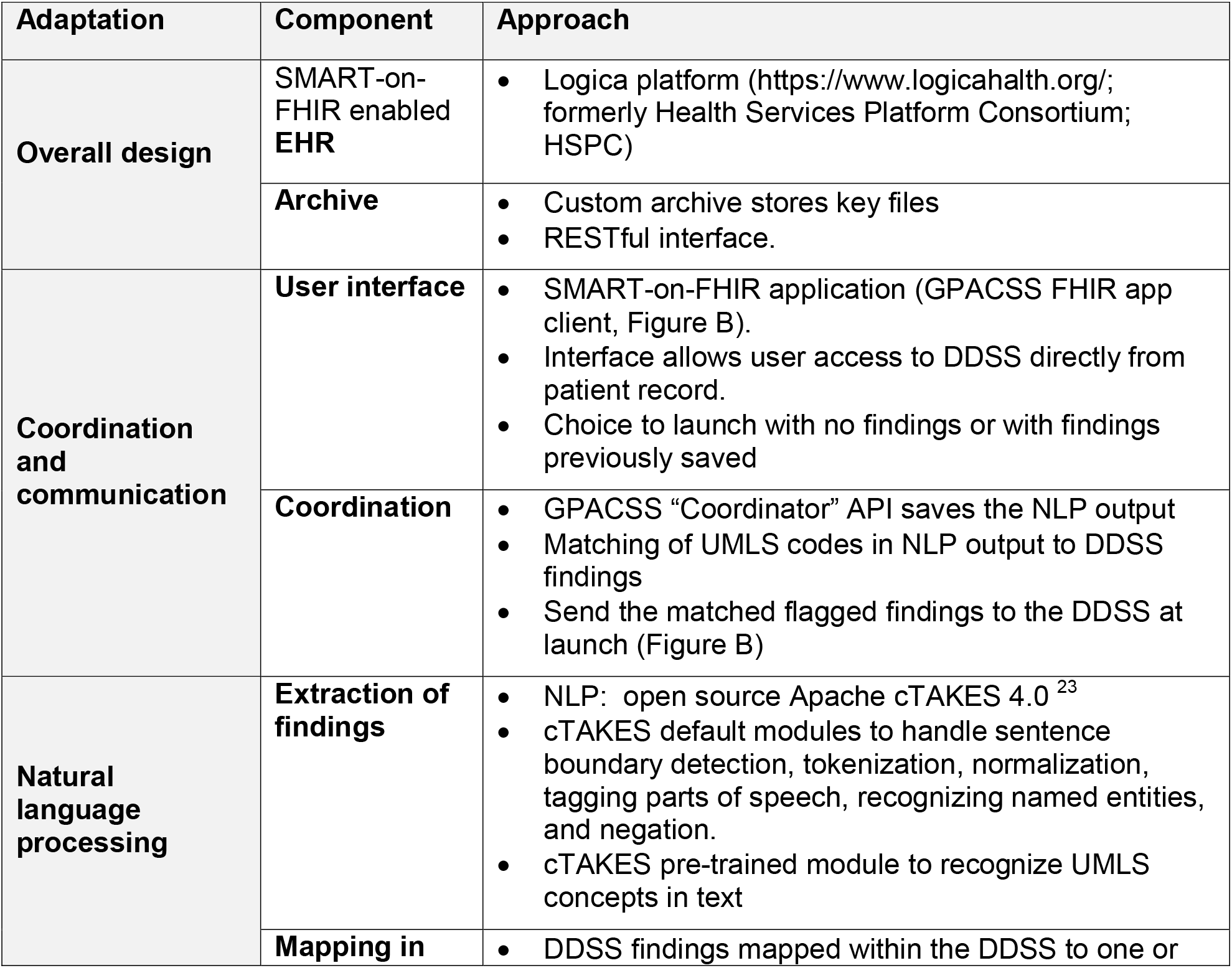
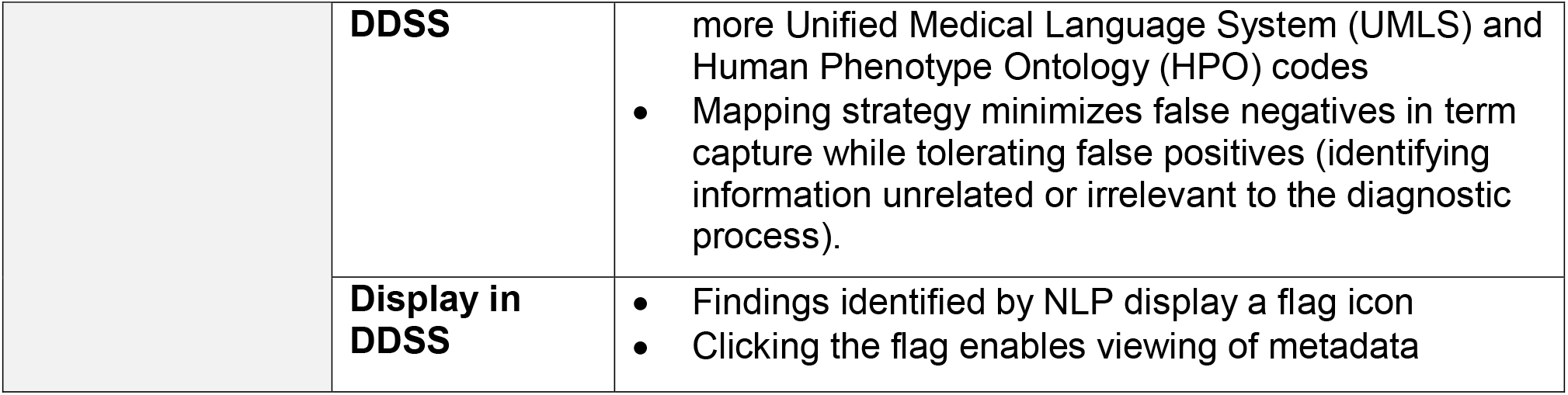
Adaptations made to existing DDSS to create GPACSS.

The architecture of the resulting prototype, called the Genotype-Phenotype Archiving and Communication System with SimulConsult (GPACSS), is shown in Figure B.

**FIGURE B:**
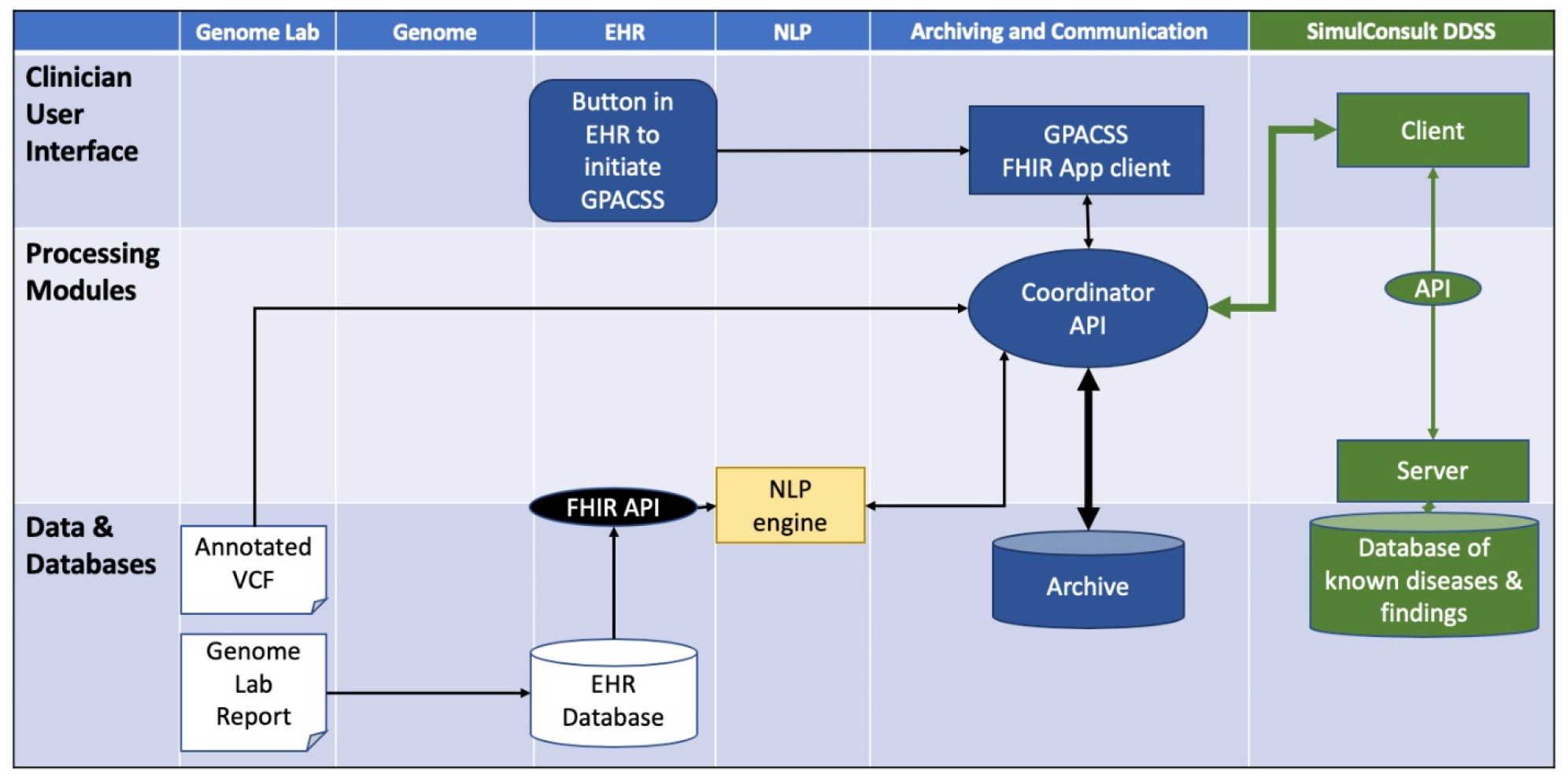
Architecture of the Genotype-Phenotype Archiving and Communication System with SimulConsult (GPACSS) The key components are the coordination / archiving system (blue), the DDSS (green) and the NLP (yellow).

Clinician review of the flagged findings created from the automated findings search using NLP is facilitated through flag icons (Figure C). Through this “machine-assisted” chart review, the clinician reviews flagged findings and decides whether and how to specify presence (with a particular onset) or absence (or omit) as shown in Figure A. The mapping of DDSS findings to multiple UMLS concepts was chosen to minimize false negatives in concept identification; relying on the user decisions about findings and the limited set of UMLS concepts to minimize false positives (Table 2).

**FIGURE C:**
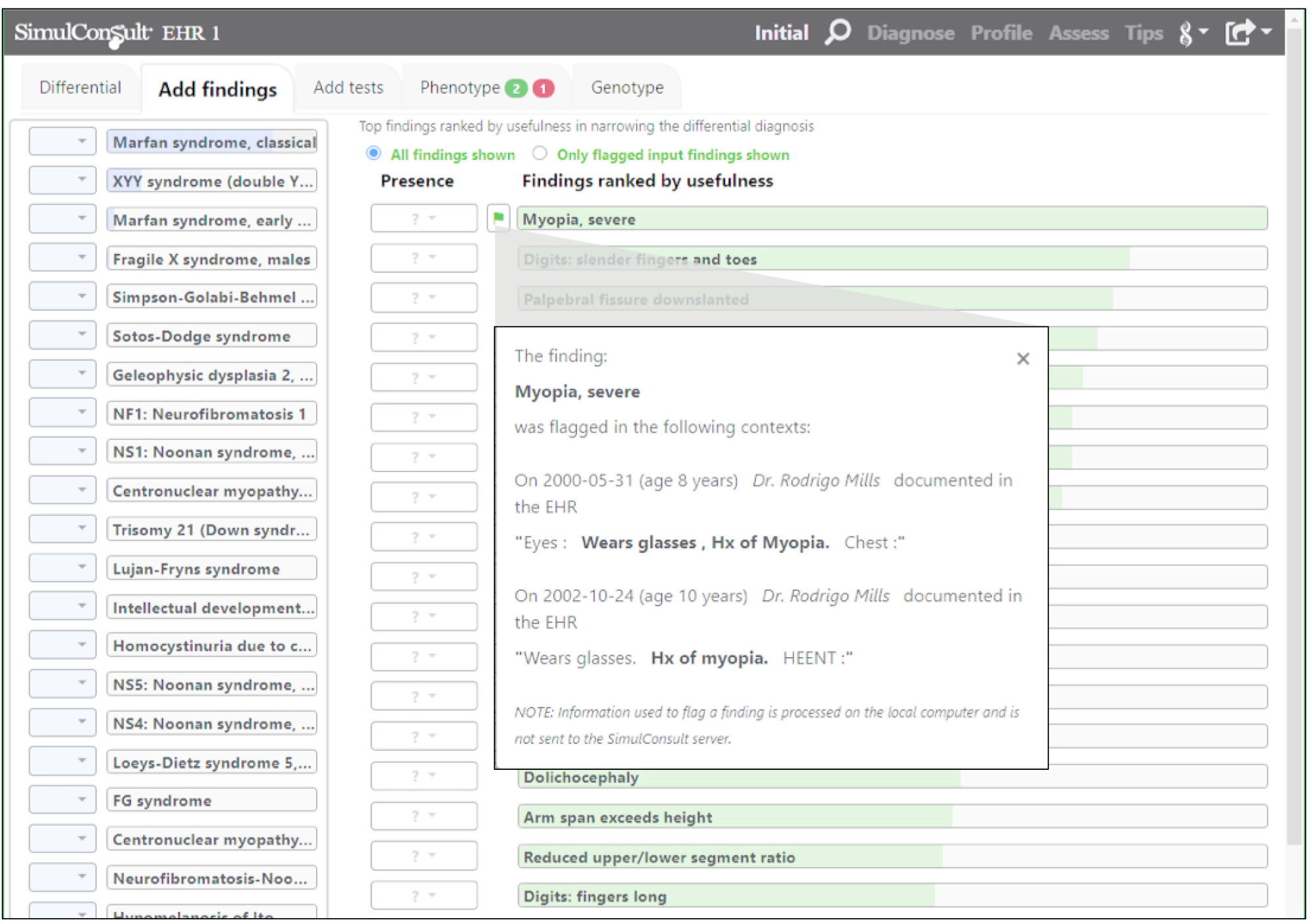
Flagged findings with EHR text display for DDSS. A finding having a flag icon indicates that information was found in the EHR. Clicking the flag shows the various mentions of the flagged finding.

**Table 2.** Solutions for Minimizing False Positives and Negatives Identified Through NLP and DDSS by Clinician Review.

| False Negative/Positive Problem | Solution Included in GPACSS |
| --- | --- |
| Minimizing false negatives on NLP flagging of findings | • Include parent and child codes (e.g. finding of intellectual disability in DDSS includes codes for developmental delay and particular types of intellectual disability) |
| Minimizing false positives through the DDSS Usefulness metric | • Use DDSS usefulness algorithm<sup>24</sup> to display flagged findings; thus prioritizing data of greater relevance and de-prioritizing data of low relevance for clinician review |
| Minimizing false positives through clinician verification | • Use flag icon to indicate findings identified through NLP (Figure C)<br>• Clinician clicks the flag icon to display information needed to assess reliability, presence or absence, and onset<br>• Information displayed from the EHR includes date of chart note, observer identity, and 3 sentences of chart note (sentence with finding plus preceding and subsequent sentence) |

### Creating simulated cases

Three cases of increasing complexity were created using real but de-identified clinical phenotypic and time course data from medical notes of Geisinger patients with known genetic diagnoses (Supplemental Table 1). Cases were selected for conditions of varying complexity yet relatively common in the context of rare disease and where diagnosis might be difficult using phenotype alone. Simulated cases were created by research assistants trained in capturing information from the EHR, supervised by a practicing Geisinger clinician certified in genetics and informatics. The three final cases were reviewed by a second Geisinger physician certified in genetics and informatics prior to user testing.

Case vignettes for the test scenarios assumed that some patient characterization was previously noted by the clinician and genomic results were now available and could be interpreted with clinical information available in the EHR (Supplemental Figure A). For the 3 cases, a total of 5 findings were used as initial information before the genomic results, with 3 (one per case) being flagged findings identified through NLP. This created a “near live”^25^ experience within the simulated EHR for user testing while limiting the expense and time of EHR integration during this preliminary phase.

### User Testing Methods

#### Participants

GPACSS is both a DDSS and communication tool to facilitate utilization of genomic and phenotypic information available in the EHR by all clinicians to improve patient care within a healthcare system. Therefore, we purposively selected primary testers from Geisinger staff representative of current end users of the genome-phenome analyzer. Because a limited number of individuals at Geisinger regularly engage in utilizing genomic information for differential diagnosis, we followed guidance recommending 3 -5 evaluators for preliminary usability testing.^26^ A group of secondary testers (inclusive of a pilot tester) with other roles in the genetic testing and interpretation process were purposively selected for potential broader utilization in the healthcare system.

#### Testing Sessions

At the beginning of each session, testers viewed a 4-minute training video (https://simulconsult.com/videogpacss) beginning from saved patient findings, then importing a VCF, and review of flagged findings to make a diagnosis and create a customizable patient-friendly RoR report.

A semi-structured interview guide was created to elucidate factors relevant to human factors in CDS design (information, interaction, interface)^1 5 27^ and organizational implementation (acceptability, perceived need, feasibility, workflow fit).^28^ We utilized a think aloud^25^ approach where testers were asked to verbalize thoughts while using the GPACSS prototype with the interviewer asking questions as needed and at key points in the testing to create a cognitive walkthrough with heuristic evaluation.^26 29^ Testers were invited via direct contact from study staff and provided a description of the study. At the beginning of each session, study staff reviewed a study information sheet and obtained verbal consent to participate. Test sessions lasted 2 hours and testers received a $100 gift card. The user testing protocol was reviewed and approved by the Geisinger IRB.

An experienced interviewer (AKR) and observer (MAW) from Geisinger worked with each tester to imagine using GPACSS for each test scenario. The interview and process were piloted with a cancer genetic counselor reviewing one test vignette. At the end of the session, testers were asked a series of study-specific questions using a 0-10 rating scale (hard to easy) to rate the overall usefulness, satisfaction, and navigation. Transcripts were created from the audio portion of each session and the computer screen was video recorded to capture tester movement through GPACSS.

### Analysis

Two Geisinger coders (MAW, JCR) viewed each user test session recording, read transcripts, and created a codebook of themes identified across sessions. Transcripts were coded and the corresponding quotes were organized into a matrix using the 3 categories of CDS components (information, interface, and interaction) identified by Miller et al,^1^ and categories of acceptability, perceived need, feasibility, and workflow fit according to Rogers’ Diffusion of Innovations in organizations constructs.^28^ Coders analyzed transcripts independently and reviewed for agreement with discrepancies resolved by the primary author.

## RESULTS

Three clinicians currently using genomic information to diagnose patients participated as primary testers: a pediatric geneticist (orders exomes daily), internal medicine physician (orders 4-5 exomes per month), and a pediatric genetic counselor. Three additional clinicians participated as secondary testers; representing broader usability within the healthcare system: the pilot tester (cancer genetic counselor), a laboratory director (conducts variant interpretation), and a laboratory genetic counselor (conducts variant analysis).

### GPACSS Usability: Human Factors of CDS Design

Overall impression of the prototype was positive. Testers raised general issues relevant to human factors in CDS design^1 5^.

#### Interface

Testers liked the flagged findings (Figure C), the contextual information for each mention in the EHR, and the rank ordering of flagged findings by usefulness. The visualization of the evolving differential diagnosis and the automated RoR report for sharing with patients and referring clinicians, including the ability to save and access this report from the EHR were also appreciated.

The interface was noted to be complex, but testers stated this was expected due to the inherent complexity of genetic diagnosis and that they anticipated a learning curve to develop proficiency. Placement, positioning, and the multiple presentation layers (text and graphics in the interface)^1^ were well liked. In particular, the “Assess diagnosis” display was noted as valuable because it made transparent the logic used by the DDSS in comparing patient findings to information about the disease. Of note, each tester interpreted differently the meaning of the graphical bars and shading, however, this did not hinder their ability to make the diagnosis, and the bar itself was appreciated as a design feature. To help with interpretation, more labeling was suggested (Table 3).

**Table 3.**
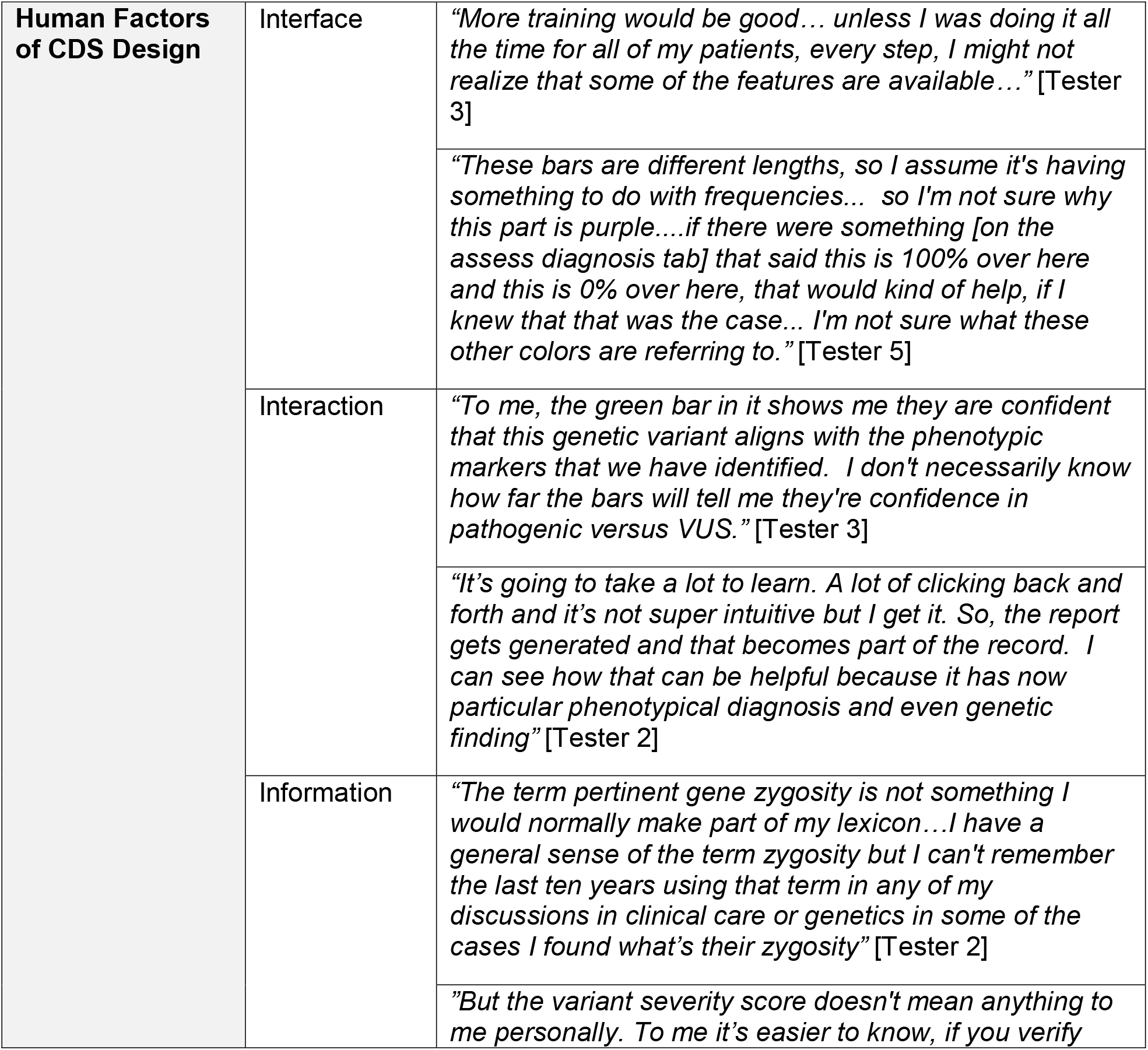
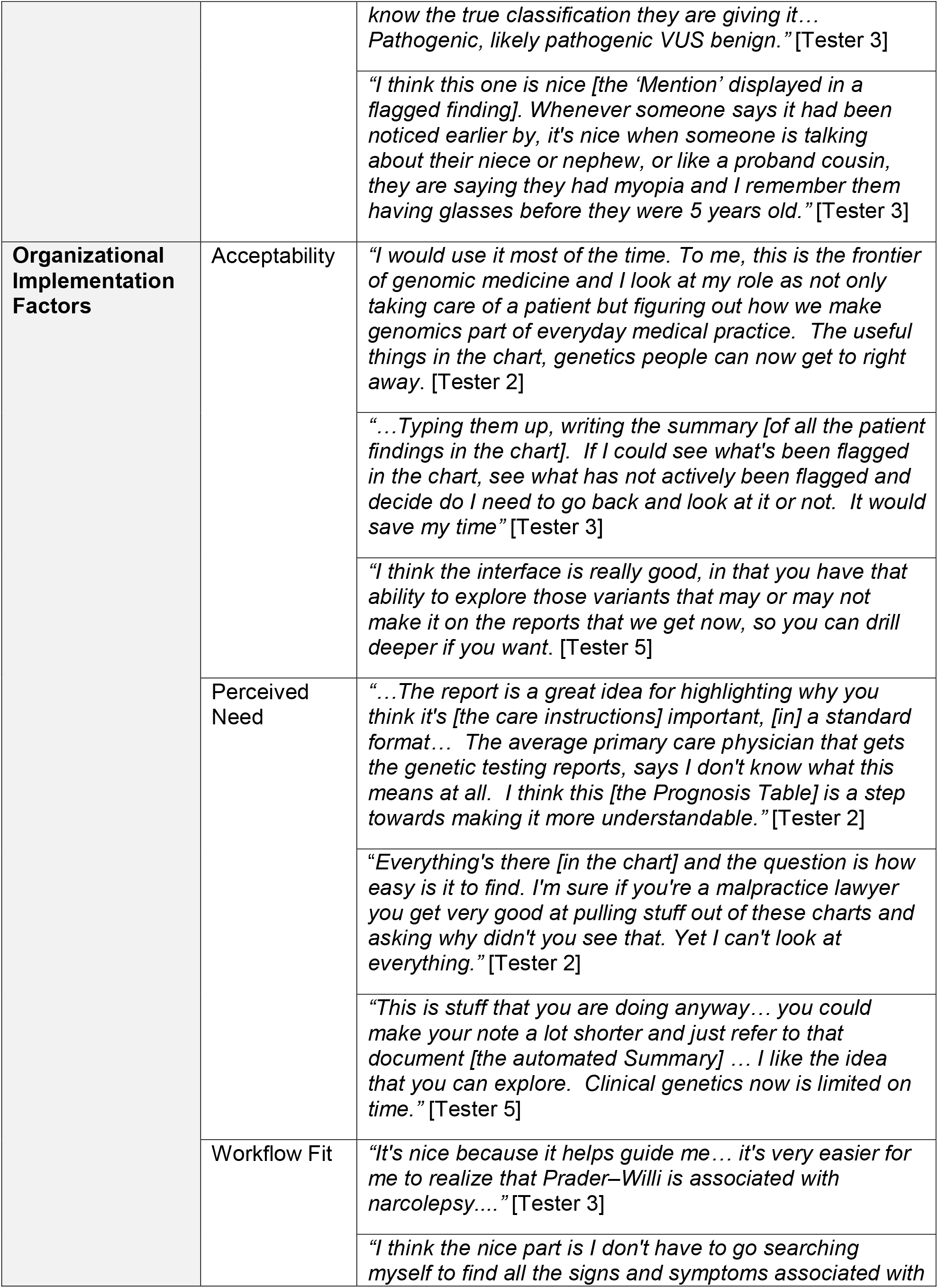
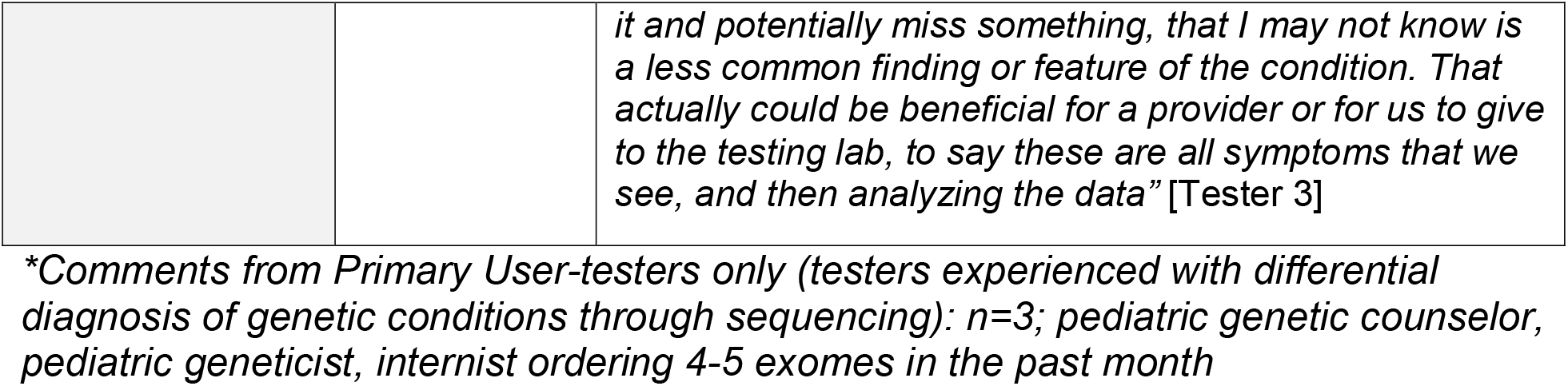
GPACSS Usability: Human Factors of CDS Design and Organizational Implementation Factors Through Tester Experiences*.

#### Interaction

Testers were thoughtful and purposeful using GPACSS. Notably, in case 3 (the most complex case), one primary tester did not immediately choose the top diagnosis offered by GPACSS. Supported by the data displayed, the tester indicated that to make a definitive diagnosis they would next evaluate for the second-ranked disease – as that condition had a test that was easy and accurate and the condition was also more treatable – indicating utilization of the DDSS as intended and consistent with clinical diagnostic decision-making.

Testers initially expressed concern around “too many clicks” and “click fatigue” but noted as they progressed through the cases that the clicking was unavoidable and necessary. For example, they saw value in taking the time to correctly specify onset information (which requires clicking and cognitive load in the DDSS), as this is part of the genetic diagnostic process. “Cognitive Load” in DDSS testing refers to additional thinking required to interact with the tool, and the general recommendation is to minimize this in CDS design.^1^ Testers who commented on the cognitive load required to review flagged findings and choose age of onset noted the cognitive load as similar to completing this task without GPACSS.

#### Information

Testers appreciated resources such as the hover feature that revealed synonyms to findings and requested even more hovers and infobuttons. Confusion over some terminology occurred, notably “zygosity” and “severity score,” when reviewing the genomic variants; as only some testers located the explanatory resource for these terms.

The fact that the EHR “Mentions” displayed in flagged findings were sometimes triggered by parent or by child concepts was noticed by all testers, and some stated the findings used in the DDSS were not as granular as they were expecting. Regardless, testers recognized and emphasized the importance of being able to review the “Mention” information from the EHR and manually adjust for any false positives and false negatives from the NLP process.

### GPACSS Usability: Organizational implementation factors

#### Acceptability

For the primary testers, satisfaction averaged 8.5 out of 10 (range 8-9.5) and navigation ease averaged 8 out of 10 (range 7.5-9). All 3 felt GPACSS would save time throughout the clinical process, with one primary tester estimating it at 30-50%. Specific value in time saved was noted for chart review by all testers.

#### Perceived need

The RoR report and detailed prognosis table^20^ generated in each scenario was highly valued for being standardized and for its ability to communicate complex genetic information to patients and other clinicians (Table 3). The RoR report was also noted as an improvement over current laboratory reports; with one tester stating it was “where the most utility would be” [Tester 4].

Testers exhibited learning and familiarity with GPACSS as they progressed through the testing session; appreciating the DDSS assistance as each vignette increased in complexity; noting *“It takes it [clinical diagnosis and diagnostic thinking] to a higher level”. [Tester 2]*. Primary testers expressed readiness to adopt the tool in clinical practice; and one (pediatric geneticist) suggested GPACSS could also serve as a differential diagnosis training tool for medical students and residents in their clinic.

Two secondary testers (lab director and variant analyst) expressed enthusiasm that GPACSS could fill a need for in-house sequencing laboratories because full EHR data would be available during sequence interpretation. These testers also hypothesized that the ability to periodically re-analyze an existing VCF in minutes using GPACSS would improve the diagnosis rate over time.

#### Workflow Fit

The three primary testers noted that the GPACSS process as tested fit with their clinical workflow diagnosing genetic conditions. As an added benefit, they described how using GPACSS also helped them learn about diseases and associated findings with which they were less familiar (Table 3).

The three secondary testers questioned GPACSS fit with a clinical genetic testing workflow in which only a report with variants labeled as to pathogenicity and association with a condition (implying a clinical diagnosis) is received from an external lab. However, they did identify value and possible workflow fit for situations with uncertainty as to the diagnosis after sequencing or where flagged findings and the usefulness ranking would allow clinicians to review the EHR with flagged findings in light of the genomic information to make the diagnosis.

## DISCUSSION

We provide preliminary evidence through user testing in a simulated real-world clinical workflow that the combination of NLP with a CDS tool optimized to support the clinical process of differential diagnosis may address the needs of those involved in this complex task. Such assessment of fit is critical if CDS is to fulfil the promise of standardizing and improving care.^1 4 5 8^

Technical issues related to the interface and interaction of CDS design were minor and fixable; as were issues with design layout. Despite initial remarks on the number of clicks and cognitive load, testers acknowledged these as necessary to the genetic diagnosis process and no different than without the DDSS. Other issues related to terminology and usability could be solved and evaluated in future usability studies through a combination of training, added infobuttons, and experience using GPACSS. Some of the technical gaps noted and additions requested by testers are addressed within GPACSS, however, the 4-minute training video was created to provide enough instruction only to facilitate user testing. These results therefore provide direction for training and ongoing reference materials for future implementation.

For CDS to be acceptable and implemented by clinicians and organizations, it must fit with the real-world workflow and must present a solution to a perceived need.^5 28^ All primary testers identified ways GPACSS added such value and fit and noted ways GPACSS filled multiple needs in their diagnostic workflow. Workflow fit was highest among primary testers but opportunities for workflow fit were described by all testers. GPACSS was also noted as acceptable for implementation by all testers regardless of individual issues identified and suggestions for technical improvements.

## LIMITATIONS

To facilitate user testing of GPACSS in the context of clinical workflow prior to full integration and implementation, simulations of the real-world were required. Because this study used the Logica EHR simulation, benefits or drawbacks of GPACSS in a production EHR could not be directly observed. Also, full annotations for the causal variants were not included in the variant table for the simulated patients limiting full assessment of the value of the DDSS in variant interpretation. This impacted the understanding of the “severity score” by all testers, as the annotation information that would have been provided for a real patient was not included for the simulated cases. Finally, the generic cTAKES NLP using the UMLS concepts found only 20 of the 30 (67%) pertinent positive concepts within the test cases that a pediatric neurologist (MMS) identified manually. This was sufficient for GPACSS to generate the correct differential diagnosis for user testing, as further enrichment of the generic NLP to improve detection and avoid false positives was out of scope for this preliminary user testing^30^. Subsequent automated search for UMLS terms for flagging and addition of a separate stage of text search enrichment for terms missed by the NLP such as “tall” improved NLP yield to 30 of 30 (100%).

This simulated EHR and user testing were a necessary first step and provide data to guide implementation of GPACSS. NLP improvements and additional beta testing within an actual EHR, in real-world clinical workflows, with real patient results, and in real-world clinical workflows will be necessary to fully assess individual user-level and organizational-level facilitators and barriers to use, implementation, and impact on clinical care. Such studies are currently in progress.

## CONCLUSIONS

This study provides preliminary evidence for the usability, workflow fit, acceptability, and implementation potential of a DDSS that includes machine-assisted chart review. Overall, responses suggest the GPACSS prototype is usable based on technical CDS and human-centered design criteria, addresses perceived clinical need, and has good fit within the real-world clinical workflow of genetic testing and diagnosis. Further development is needed to improve usability for multiple clinical stakeholders and organizational implementation.

### Contributors

All authors reviewed the final manuscript for submission and contributed to the study as follows:

AKR contributed to the study design, conducted user testing sessions, and led the analysis, interpretation, and writing of manuscript.

NAW contributed to the study design, created the simulated clinical environment and the de-identified patient records, contributed to the NLP testing and adaptation of the DDSS and manuscript revisions.

TNP was the lead developer on the NLP and the integration with the SimulConsult API. CJ, TJ, TNP, and JP contributed to NLP testing and creation of the simulated clinical environment and manuscript revisions

MAW contributed to project management, user testing recruitment, qualitative data analysis and interpretation and review of manuscript.

JCR contributed to project management, IRB applications, NLP testing and creation of the simulated clinical environment, and manuscript writing

MSW contributed to study design, simulated clinical environment, NLP testing and adaptation of the DDSS, user testing design and recruitment, data analysis, interpretation and manuscript writing

LKF contributed to project management, study design, and manuscript revisions

PNR assisted in the tagging of DDSS findings with HPO codes and created the new codes when needed.

MMS designed and implemented the adaptation of the clinical decision support tool to interact with the NLP, designed and implemented the strategy for HPO and UMLS coding, did the manual chart review to identify findings that should have been recognized by NLP, and contributed to the study design, and manuscript revision

## Supporting information

Supplemental interview guide

## Data Availability

study data are qualitative interview transcripts and are not made publicly available, however, authors may be contacted for information.

## FUNDING

This study was supported by the National Human Genome Research Institute of the National Institutes of Health under the Small Business Innovation Research (SBIR) Program, Award Number 1R43HG010322-01 (principal investigator: MMS). The content is solely the responsibility of the authors and does not necessarily represent the official views of the National Institutes of Health.

### PATIENT CONSENT FOR PUBLICATION

not required

### ETHICS APPROVAL

This study was reviewed and approved by the Geisinger Institutional Review Board (IRB)

### PROVENANCE AND PEER REVIEW

Not commissioned; externally peer reviewed

### DATA AVAILABILITY STATEMENT

Data are available upon reasonable request.

