## Supplemental interview guide for "User Testing of a Diagnostic Decision Support System with Machine-assisted Chart Review to Facilitate Clinical Genomic Diagnosis"

GPACSS User Testing Interview Guide

**Consent/Recording:**

*Thank you for agreeing to participate in our testing of the Genome-Phenome Archiving and Communication System (GPACS). Our goal today is to have you simulate a clinical process for each of 3 case scenarios - going from assessment to diagnosis after genetic sequencing results. We will be recording how you use GPACS in each case, how easy/difficult it is to find what you are looking for, what works/not for you, and what your thoughts are on how helpful it is in this process. I will also be asking you to “think aloud” as you are going through this – we will be recording your thoughts and clicks, and I will be asking general and specific questions throughout.*

*You can stop participating at any time. At the end you will receive a gift card.*

**Start recording**

**Introduction and video instruction:**

*I’m going to present you with 3 clinical vignettes about cases where you might order genome/exome sequencing for the patient. After the first vignette I will show a short video that will orient you to the Genome-Phenome analyzer and how to use it to help you facilitate the diagnostic process.*

*I’ll be asking questions as you go through each vignette, and we can watch the video again if needed.*

*This video will orient you to the GPACS tool as it would look in the EHR once you open with findings and start the process of reviewing the case and the genetic results. The tool finds many, but not all findings in the EHR. We will start the first vignette after you watch the video:*

**launch user testing orientation video**

***For each vignette:*** *Imagine you have returned the patient’s chart and have opened Simulconsult with prior findings. Look around and see if you can see the findings and import the genome. Then let’s do like the video and see the findings, review the chart, and create the report*

Tester process for each vignette:

1. Open with pre-loaded findings and any family history
2. Import genomic results
3. Review flagged findings and “Mentions”
4. Record pertinent negatives
5. Make diagnosis and create report

**Launch GPACS Logica sandbox**

**Vignette 1:**

*You have seen a 27 year old male with pectus excavatum and tall stature. As part of your clinical process you have ordered genomic sequencing for this patient. After the genetic information is available, you return to the patient’s chart to review the genetic information in light of the patient’s other medical information. You launch the GPACSS system and click in as a permitted user, choose a patient, and launch SimulConsult with the prior findings already entered*

**Vignette 2:**

*You have seen a 21 year old female with seizures who sometimes has some hand wringing. As part of your clinical process you have ordered genomic sequencing for this patient. After the genetic information is available, you return to the patient’s chart to review the genetic information in light of the patient’s other medical information. You open an encounter and launch the GPACSS system with Your prior findings*

**Vignette 3:**

*You have seen a 14 year old female with seizures and a small head. As part of your clinical process you have ordered genomic sequencing for this patient. After the genetic information is available, you return to the patient’s chart to review the genetic information in light of the patient’s other medical information. You open an encounter and launch the GPACSS system with Your prior findings*

**Interview Questions during each case scenario (asked as appropriate in vignettes):**

**General question throughout: Why did you look there? Do that? Think of that? Click that? You seem to be clicking around – are you looking for something specific?**

- How does the GPACSS process fit with what would normally do in this process?
  - How does GPACSS help/not?
  - What do the numbers/severity score mean to you?
  - What does the list/shading/differentials/graphics mean to you (or do for you)? **(at each screen)**
    - How do they help (or not)?
- How does GPACSS help you locate info in chart and decide whether it is relevant? How does the ordering of findings by usefulness help you focus on relevant information?
- What questions does using GPACSS bring up for you as you’re using it?
  - Thoughts on the experience so far?
- What are your thoughts about the report? How would you use it? What are your questions about it?
- What would you do next in this process/case?
- Is there anything in this process you WOULDN’T use for this case? (why?)
- How did using GPACSS for this case compare with your typical process?
  - Was GPACSS intuitive?
  - What did you think about the filtering/flagging?
    - How did it make the process easier/harder?
    - What questions did you have about it as you were using it for this case?
    - Would you like more text before or after the text in bold of the mentions of a finding in the chart?
    - Would you like a hyperlink to open up the whole note in which a mention was found?
  - How difficult / easy? What needs to change?
  - Does this process fit with workflow? How/not?
- What would you do next with this case after reviewing the exome results?

**CLOSING QUESTIONS:**

1. How satisfied are you with whole GPACS process? (0-10 scale from “not at all” to “extremely satisfied”)
   - Explain
2. How often do you think you would you use it when receiving exome results? (0-10 scale from never to always)
   - Explain
3. How confident were you with the GPACSS diagnostic support process? (0-10 scale from “not at all” to “extremely confident”)
   - Explain
4. How comfortable were you with the flagged findings and chart context filters? (0-10 scale from “not at all” to “extremely comfortable”)
5. How do you see yourself using this for your patients regularly?
   - What works for you to use this regularly
   - What needs changing for you to use it regularly?
6. How does GPACSS improve/not the process for you?
7. Workflow thoughts? Where to implement? What needed to implement?
8. What sort of training would you need to use this regularly?
9. Is this something you would integrate into practice? Explain.
10. How would you use it?

- How would colleagues use it?
- Other feedback?

**DEMOGRAPHIC QUESTIONS:**

1. How long have you been at Geisinger?
2. How long have you been at your current position?
3. Current department and clinical role?
4. How often do you order exomes and report results?
