## Supplemental Table 1 for "User Testing of a Diagnostic Decision Support System with Machine-assisted Chart Review to Facilitate Clinical Genomic Diagnosis"

**Supplemental Table 1. Creating and processing simulated patients* in GPACSS for user testing in simulated real-time clinical workflow**

| **Simulation aspect** | **Process used in simulation** |
| --- | --- |
| Phenotype | All findings discussed in the original Geisinger EHR text were retained but notes were manually rewritten by changing narrative to remove identifying information and prevent re-identification, shifting dates while maintaining chronological relationships, and shifting numeric values while maintaining appropriate range to retain clinical meaning (i.e. laboratory values for “high-abnormal” range were maintained in that value range). |
| Genotype | Genomic data was simulated by adding the known causal variant to variant tables generated from publicly available trios from the 1000 Genomes Project to prevent identification of a real person. However, the known causal variant was not given key annotations such as functional and conservation scores that are used in the DDSS’s explanation of variant and zygosity severity scores. |
| Processing | Resulting simulated patient data for 3 simulated patients was loaded into the Logica platform for user testing and run through the cTAKES Clinical Pipeline using default settings. |

**3 simulated patients of escalating complexity with notes and clinical findings were created based on 3 real Geisinger patients with known genetic disorders. Two others were used in earlier testing and the demo video (*<https://simulconsult.com/videogpacss>*).*
