## Supplemental Figure 1 for "User Testing of a Diagnostic Decision Support System with Machine-assisted Chart Review to Facilitate Clinical Genomic Diagnosis"

Supplemental figure A

SUPPLEMENTARY Figure A: SimulConsult interface showing Gene zygosities found in the genomic analysis

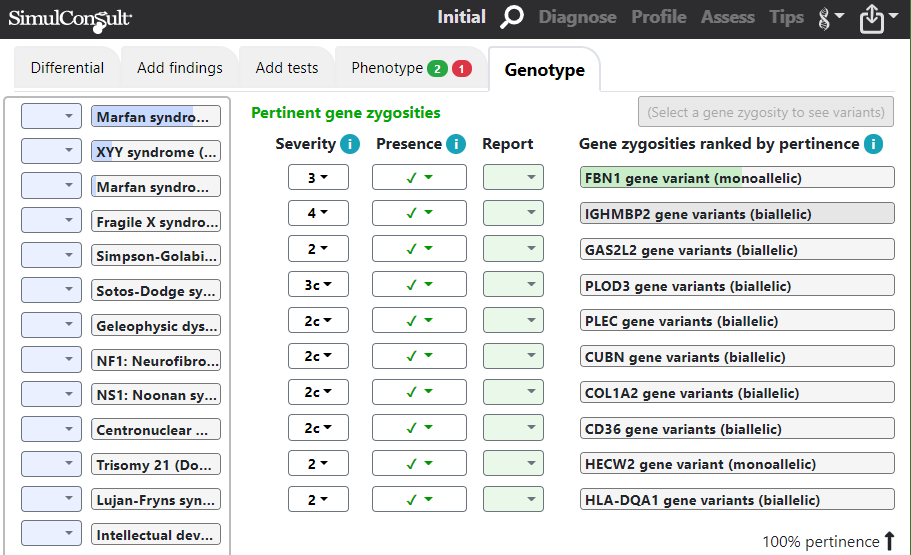
